# Evolving impact of the COVID-19 pandemic in chronic dialysis recipients over the course of pandemic waves and COVID-19 vaccination rollout: a French national study

**DOI:** 10.1101/2024.04.05.24305315

**Authors:** Elhadji Leye, Khalil El Karoui, Tristan Delory, Maude Espagnacq, Myriam Khlat, Sophie Le Coeur, Nathanaël Lapidus, Gilles Hejblum, the COVID-HOSP working group

## Abstract

**Background:** This observational study aims to assess the impact of the pandemic on the evolving of kidney transplantations, survival, and vaccination in chronic dialysis recipients (CDR) over the COVID-19 pandemic subperiods.

**Methods:** Using the French national health claims database, incident persons with end stage kidney disease in years 2015 to 2021 treated with dialysis were followed-up until December 31, 2022. Kidney transplantion and survival over pandemic subperiods versus the prepandemic period were investigated using longitudinal models with time-dependent covariates. Moreover, the impact of cumulative doses of COVID-19 vaccine on hospitalization and survival were compared between CDR and matched-control individuals.

**Findings:** Follow-up of the 71,583 CDR and 143,166 controls totalized 639,341 person-years (CDR: 184,909; controls: 454,432). The likelihood of receiving a kidney transplant decreased during all pandemic subperiods except one. Mortality in CDR increased during the 3 wave subperiods (hazard ratio (HR [95% confidence interval]): 1.19 [1.13–1.27], 1.19 [1.15–1.23], and 1.12 [1.07–1.17], respectively). While vaccine coverage declined with each booster dose, receiving such doses was associated with lower risks of COVID-19-related hospitalization (0.66 [0.56–0.77], 0.83 [0.72–0.94] for 1^st^ booster versus 2^nd^ dose and 2^nd^ booster versus 1^st^ booster, respectively) and death (corresponding HR: 0.55 [0.51–0.59], 0.88 [0.83–0.95]). Evolving patterns in mortality and vaccination outcomes were similar in CDR and controls.

**Interpretation:** The impact of the pandemic in CDR was not specific of the kidney disease *per se*. Study results also suggest future research aimed at increasing adherence to vaccine booster doses.

**Funding:** Initiative Économie de la Santé de Sorbonne Université (Idex Sorbonne Université, programmes Investissements d’Avenir) ; Ministère de la Solidarité et de la Santé (PREPS 20-0163).

## 1. Introduction

The COVID-19 pandemic rapidly plunged the world into an unprecedented systemic crisis in 2020. On 30 January, 2020 the World Health Organization (WHO) declared COVID-19 a global public health emergency,^1^ leading to multiple lockdowns worldwide, including France, for mitigating the spread of the virus.^2^ Early studies highlighted a higher risk of hospitalization and mortality from COVID-19 in persons under maintenance dialysis due to an end-stage kidney disease (ESKD).^3,4^ After the first COVID-19 vaccination campaigns, several studies reported lower responses to vaccines and a faster waning of immunity in chronic dialysis recipients (CDR) as compared to the general population,^5,6^ and the requirement of additional booster doses 4 to 6 months after prime-boost vaccination has been advocated for protecting as much as possible this more vulnerable population.^7^ In addition, adherence/hesitancy of CDR to such booster campaigns remains insufficiently documented in the literature. Indeed, a recent systematic review examining the relationships between Health Belief Model constructs and COVID-19 vaccination intention included 109 studies but only 13 articles explored booster vaccines,^8^ and only 1 of these 13 articles explored the booster vaccine hesitancy in hemodialysis patients.^8,9^ Such findings led us to undertake investigations on vaccination coverage for booster doses. Another challenging consequence of the COVID-19 pandemic for CDR was the reduction in kidney transplant activity. An overall decrease of 8,560 kidney transplants across 22 countries was reported in an international study, including a decrease of 1041 (−34%) kidney transplants in France between February 29, 2020 and December 31, 2020.^10^ To our knowledge, despite the well-established impact of COVID-19-related hospitalizations on CDR survival,^3,4^ a detailed analysis of the likelihood to receive a kidney transplant over the course of the pandemic has not been reported to date.

In order to assess the global impact of COVID-19 pandemic on CDR, we undertook a large comprehensive study covering the entire span of COVID-19 crisis. Adopting the perspective of a national level in France, the study reported here had several objectives. Firstly, it aimed to characterize the evolving risk of mortality and likelihood to receive kidney transplant in CDR during the different waves and interwave subperiods of the pandemic. Secondly, it aimed to investigate the indirect impact of the pandemic on the survival and transplantation access in CDR. Assessing this indirect impact required examining the direct effects of COVID-19 hospitalizations and vaccinations among CDR. Lastly, analyses on appropriate control subpopulations were simultaneously conducted in order to investigate the extent to which estimates relating to survival and vaccination in CDR were specific of this particular subpopulation.

## 2. Methods

This observational study was conducted according to STROBE guidelines.^11^

### 2.1 Data Sources

The French “Système National des Données de Santé” (SNDS) was the unique source of data used in the study. SNDS is a national claim database designed for reimbursing care throughout the French Health Insurance System covering nearly 100% of the population receiving heathcare.^12^ With the growing interest of the scientific medical community in real-world data from large administrative healthcare databases, the SNDS has been previously described and used for pharmacoepidemiologic studies,^13–15^ including studies on COVID-19.^16,17^

### 2.2 Patients included in the study

ESKD incident persons in the SNDS were identified using G10 mapping,^18^ as previously described in a study investigating the direct and indirect impact of the pandemic on the survival of kidney transplant recipient.^19^ Only individuals aged 18 years or more at the time of incidence and whose first care related to ESKD was dialysis (peritoneal or hemodialysis) were included in the study. This restriction to incident persons with an ESKD treated by dialyses is an essential feature of the study, and was adopted in order to guarantee that the age of the disease was duly documented for each individual included in the study. Each CDR included in this study was matched at the beginning of his/her follow-up with two controls persons identified in the SNDS without ESKD but with the following identical characteristics (exact matching): sex, age, history of diabetes, history of chronic cardiovascular diseases, and region of residence.

### 2.2 Prepandemic and pandemic periods, wave and interwave subperiods, vaccination roll-out

The number of COVID-19 reported cases in France rose from 100 to 1000 between February 29 and March 8, and a lockdown started on March 17. Therefore, the study prepandemic period was defined as the period ranging from January 1, 2015 to February 29, 2020 and the pandemic period began on March 1, 2020. The pandemic period was censored on December 31, 2022 considered as the end date of the study. In order to investigate potential patterns along the course of the pandemic period, the pandemic period was divided into 6 subperiods according to the frequency of COVID-19-related hospitalization admissions in the SNDS: 3 wave subperiods, (from March 01 to May 18, 2020 for the first wave subperiod, from September 07, 2020 to May 10, 2021 for the second wave subperiod, and from November 21, 2021 to April 25, 2022 for the third wave subperiod) and 3 interwave subperiods (from May 19, to September 6, 2020 for the first interwave, from May 11, to November 10, 2021 for the second interwave, and from April 26 to December 31, 2022, for the third interwave subperiod). In France, the roll-out of COVID-19 vaccination began on December 27, 2020.

### 2.3 Statistical analyses

The ESKD incidence date was considered as the initial time (t_0_) of the corresponding individual follow-up. The individual end of follow-up was censored at December 31, 2022, end date of the study. Figure 1 describes the follow-up of typical persons included in the study. Importantly, the present study investigated CDR because in a recent study, we already detailed the impact of the pandemic on kidney transplant recipients.^19^ Therefore, persons entering ESKD status with a pre-emptive transplantation event were not considered in the present study, and CDR follow-up was ended in case of kidney transplant or death.

**Figure 1.**
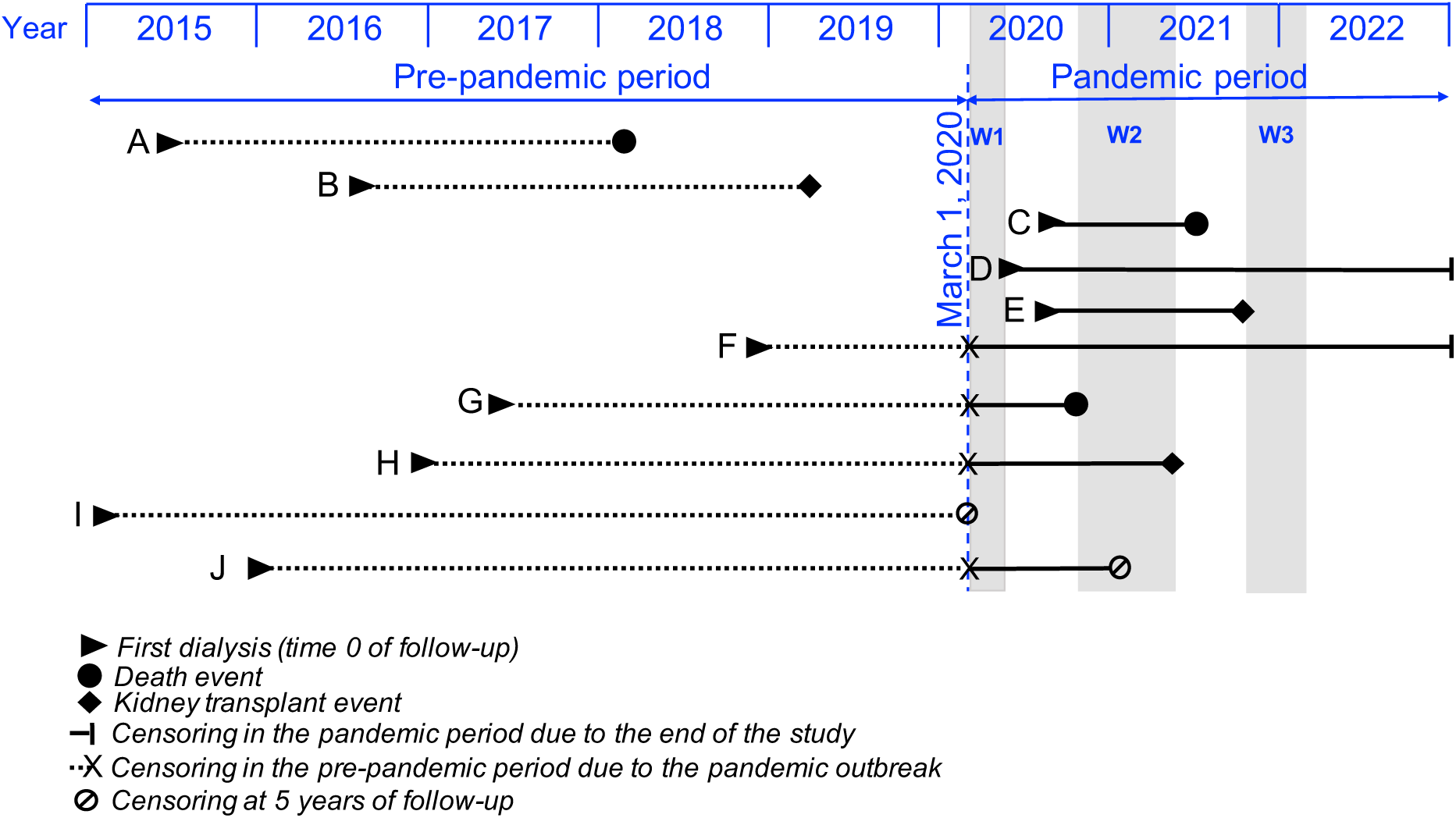
Graphic examples of some individual follow-up along the prepandemic and pandemic periods. (A-E) Whole follow-up of the person in either the prepandemic (person A or B) or the pandemic (person C, D, or E) period; (F-H, J) whenever a person became an incident chronic dialysis recipient during the prepandemic period and survived until the beginning of the pandemic period, this person left the prepandemic follow-up group and entered the pandemic follow-up group when the pandemic period began; (I and J) follow-up of any person included in the study has been censored at 5 years since there would not be any control observation of a person with a follow-up duration greater than 5 years and 2 months

Investigations of the evolving impact of the COVID-19 pandemic in CDR according to wave sub-periods required to consider additional covariates that we deemed important to simultaneously study: COVID-19 as the principal or related code (International Classification of Diseases, 10^th^ revision) for hospitalization which was considered as a proxy for a severe COVID-19 episode, age, sex, diabetes and chronic cardiovascular diseases (defined in the G10 mapping of the SNDS as the presence of at least one of the following features: stroke sequelae, chronic heart failure, coronary diseases, peripheral arterial obliterative disease), were also considered as additional covariates. The age variable was transformed into an ordinal variable handled according to the following breakdown: 18–44, 45–54, 55– 64, 65–74 (reference), 75–84, and ≥85 years. COVID-19-related hospitalization was also considered as an ordinal variable with 2 modalities: 0 and ≥1 hospitalizations. Some covariates might have varied during the longitudinal follow-up of the individuals, and accordingly, the following variables were appropriately handled as time-dependent variables in the analyses: period (pandemic versus prepandemic), pandemic subperiods (1^st^ wave, 1^st^ interwave, 2^nd^ wave, 2^nd^ interwave, 3^rd^ wave, and 3^rd^ interwave), COVID-19-related hospitalization, and COVID-19 vaccination (0 or 1 dose, 2 doses, 3 doses–first booster dose is further referred to as a third dose, and 4 doses–second booster dose is futher referred to as a fourth dose). There was no missing variable value in our database queries, and therefore, analyses assumed the absence of missing data.

Survival models with time-dependent covariates were used to assess how the COVID-19 pandemic affected both the likelihood to receive a kidney transplant and the risk of death in CDR. Competing risks of death and kidney transplantation were handled using Cox cause-specific regression models. Furthermore, study investigations extended to analyzing the association, between each additional dose of COVID-19 vaccine and two outcomes: COVID-19-related hospitalization and survival. Such investigations involved comparing individuals who received a given dose with those who had received the previous dose, using Cox cause-specific regression models with time-dependent covariates accounting for changes in vaccination status over time. As the choice of cause-specific models to handle competing risks in the presence of time-dependent covariates is debatable, we conducted sensitivity analyses using the Fine and Gray model instead. ^20–22^ Analyses were performed using R version 4.1.2. A p value < 0.05 was considered statistically significant.

### 2.4 Ethics statement

The SNDS is a set of strictly anonymous databases comprising all mandatory national health insurance reimbursement data. Since June 30, 2021 INSERM has direct access to the SNDS. This permanent access is provided according French Decree No. 2016-1871 of December 26, 2016 relating to the processing of personal SNDS data and French law articles Art. R. 1461-1325 and 14. This study was declared prior to its initiation to the CépiDc-INSERM registry of studies requiring the use of the SNDS. In accordance with national legislation and EU General Data Protection Regulation, written informed consent for participation was not required for this study.

## 3. Results

Figure 2 illustrates the study profile: considering the whole population of adult persons living with ESKD in France each year from 2015 to 2021 (prevalent cases), the selection process for incident ESKD cases undergoing dialysis during this period resulted in a total of 71,583 CDR and 143,166 matched-control individuals (2 matched-control individuals per CDR) with a total follow-up of 639,341 person-years, (184,909 for CDR and 454,432 for matched-control individuals). Table 1 presents the characteristics of the CDR included in the study at the time of incidence. Nearly 83% of the recipients were at least 55-years old, 65% were males, and almost 68% presented with diabetes, a chronic cardiovascular disease, or both. Control individuals had identical baseline characteristics to those of CDR as determined through exact matching. Considering the 98,444 person-years of cumulated follow-up in the CDR during the prepandemic period, 14,455 deaths (1468 per 10,000 person-years) and 5,853 kidney transplants (595 per 10,000 person-years) occurred. Considering the 88,603 person-years of cumulated follow-up in the CDR during the pandemic period, 13,932 deaths (1573 per 10,000 person-years) and 4172 kidney transplants (471 per 10,000 person-years) occurred. In comparison, considering the matched-control individuals, 10,898 deaths occurred during the 231,632 person-years of cumulated follow-up in the prepandemic period, (470 deaths per 10,000 person-years) and 11,387 deaths occurred during the 222,760 person-years of cumulated follow-up in the pandemic period (511 deaths per 10,000 person-years).

**Figure 2.**
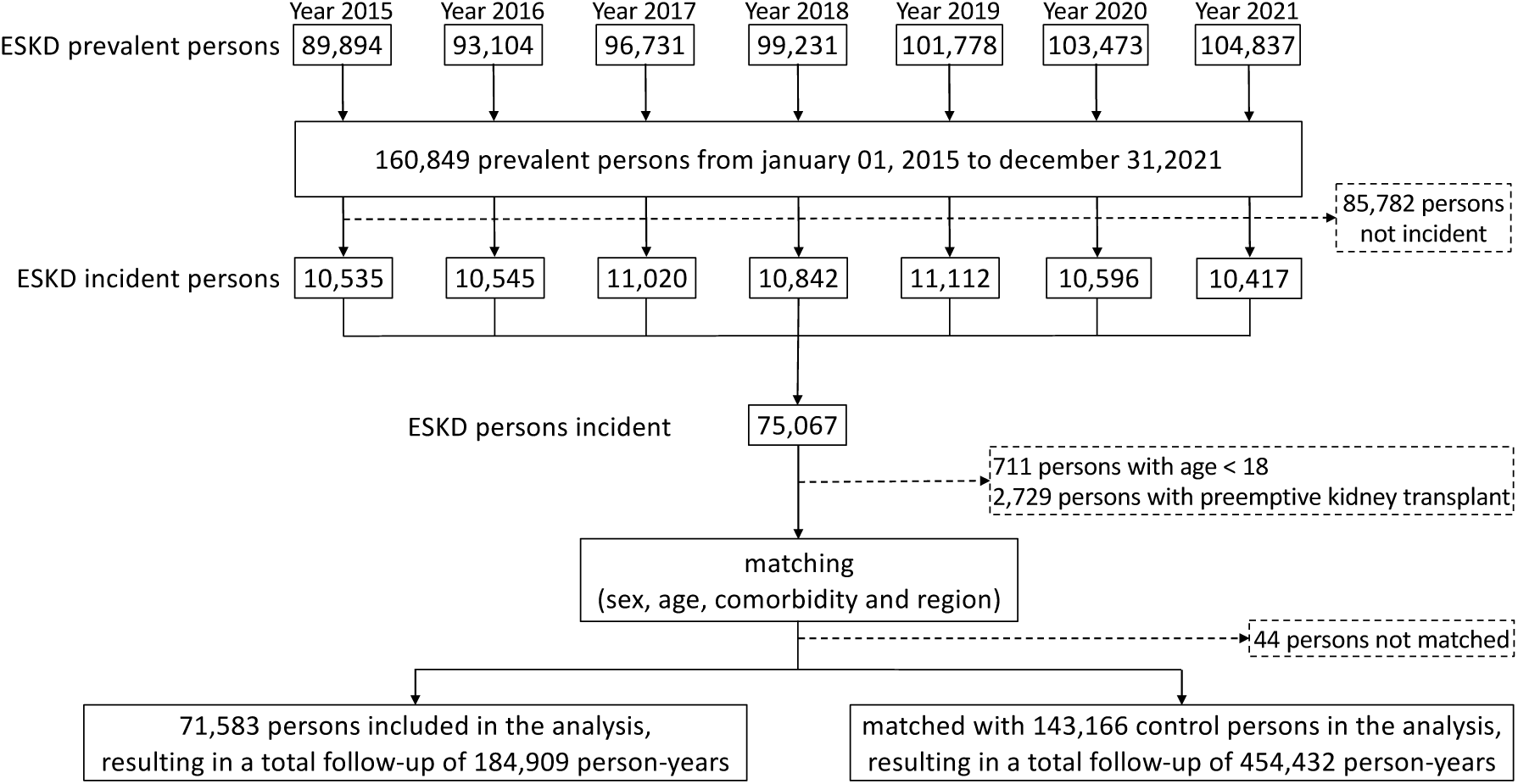
Study flow chart.

**Table 1.** Baseline characteristics of the 71,583 study CDR at the date of dialysis onset.

| **Variable** | **Variable Value** | **All CDR (n=71,583)** | **CDR with an incidence date during the prepandemic period (n=53,405)** | **CDR with an incidence date during the pandemic period (n=18,178)** | **p value** |
| --- | --- | --- | --- | --- | --- |
| **Age group** |  |  |  |  | **<0.01** |
|  | **18–44** | 5,591 (7.8%) | 4,153 (7.8%) | 1,438 (7.9%) |  |
|  | **45–54** | 6,069 (8.5%) | 4,545 (8.5%) | 1,584 (8.4%) |  |
|  | **55–64** | 11,218 (15.7%) | 8,404 (15.7%) | 2,814 (15.5%) |  |
|  | **65–74** | 19,889 (27.2%) | 14,548 (27.2%) | 5,341 (29.4%) |  |
|  | **75–84** | 20,082 (28.3%) | 15,126 (28.3%) | 4,956 (27.3%) |  |
|  | **≥ 85** | 8,734 (12.4%) | 6,629 (12.4%) | 2,105 (11.6%) |  |
| **Sex** |  |  |  |  | **0.01** |
|  | **Male** | 46,099 (64.1%) | 34,252 (64.1%) | 11,847 (64.4%) |  |
|  | **Female** | 25,484 (35.9%) | 19,153 (35.9%) | 6,331 (35.6%) |  |
| **Diabetes** |  |  |  |  | **<0.01** |
|  | **No** | 38,244 (53.9%) | 28,810 (53.9%) | 9,434 (51.9%) |  |
|  | **Yes** | 33,339 (46.1%) | 24,595 (46.1%) | 8,744 (48.1%) |  |
| **Chronic cardiovascular disease** |  |  |  |  | **0.03** |
|  | **No** | 36,060 (50.1%) | 26,774 (50.1%) | 9,286 (51.1%) |  |
|  | **Yes** | 35,523 (49.9%) | 26,631 (49.9%) | 8,892 (48.9%) |  |

Abbreviation used: CDR, chronic dialysis recipients

### 3.1 Transplantation

The decrease in kidney transplantation activity in France during the pandemic period is detailed in Figure S1 (see Supplementary Appendix). In parallel, considering the whole pandemic period, the likelihood of CDR to receive a kidney transplant decreased by 20%, as compared to the prepandemic period (hazard ratio (HR) [95% confidence interval] at 0.72 [0.70–0.76], see Table 2, model 1). Moreover, most of the corresponding CDR were without a history of COVID-19-related hospitalization, but CDR having experienced a COVID-19-related hospitalization had a 29% additional decrease in the likelihood to receive a kidney transplant (HR = 0.71 [0.59–0.85], see Table 2 model 2). In the end, considering the evolving HR for receiving a kidney transplant according to the wave and interwave pandemic subperiods as compared to the prepandemic period (Table 2, model 3), a decrease occurred during all the waves and interwaves, except the 3^rd^ interwave subperiod, with the highest decrease observed during the first wave subperiod (HR = 0.16 [0.12–0.20]).

**Table 2.** Multivariable analysis of kidney transplantation events in the 71,583 CDR during the prepandemic and pandemic periods (cause-specific Cox models).

| Model | Variable | Kidney transplantations (n) / person-years (n) | HR [95% CI] | p value |
| --- | --- | --- | --- | --- |
| Model 1 | Prepandemic period | 5,853 / 98,444 | 1 (reference) |  |
|  | Pandemic period | 4,172 / 86,465 | 0.72 [0.70–0.76] | <0.01 |
| Model 2 | Prepandemic period | 5,853 / 98,444 | 1 (reference) |  |
|  | Pandemic period, no history of COVID-19-related hospitalization | 4,060 / 82,804 | 0.73 [0.70–0.76] | <0.01 |
|  | Pandemic period, ≥1 COVID-19-related hospitalization | 112 / 3,661 | 0.71 [0.59–0.85] | <0.01 |
| Model 3 | Prepandemic period | 5,853 / 98,444 | 1 (reference) |  |
|  | 1<sup>st</sup> wave | 67 / 6,837 | 0.16 [0.12–0.20] | <0.01 |
|  | 1<sup>st</sup> interwave | 505 / 9,892 | 0.80 [0.73–0.88] | <0.01 |
|  | 2<sup>nd</sup> wave | 1,024 / 21,604 | 0.73 [0.69–0.79] | <0.01 |
|  | 2<sup>nd</sup> interwave | 874 / 17,503 | 0.77 [0.71–0.82] | <0.01 |
|  | 3<sup>rd</sup> wave | 673 / 13,075 | 0.76 [0.70–0.82] | <0.01 |
|  | 3<sup>rd</sup> interwave | 1,029 / 17,553 | 0.81 [0.76–0.87] | <0.01 |

All HR shown correspond to cause-specific Cox regressions also adjusted for the following covariates: sex, age category, diabetes, and cardiovascular disease. Abbreviations used: CI, confidence interval; CDR, chronic dialysis recipients; HR, hazard ratio.

### 3.2 Mortality

The risk of death in CDR increased during the pandemic period as compared to that in the prepandemic period: HR = 1.08 [1.05-1.10] (Table 3, Model 4). However, Model 5 further indicates that compared to the prepandemic period, the risk of death in CDR without any history of a COVID-19-related hospitalization was similar (HR = 0.98 [0.95–1.00]), while the risk of death was dramatically higher in those who had experienced a COVID-19-related hospitalization (HR = 3.48 [3.32–3.65]). Regarding the mortality trends along the pandemic subperiods (Table 3, model 6), each of the three wave subperiods was associated with a higher risk of death as compared to the prepandemic period, with respective increments of 19% (HR = 1.19 [1.13–1.27]), 19% (HR =1.19 [1.15–1.23]), and 12% (HR = 1.12 [1.07– 1.17]), while interwave subperiods were not associated with different risks of death, except for the first interwave (HR = 0.93 [0.88–0.97], 0.98 [0.94–1.02], 1.03 [1.98–1.07] for 1st, 2nd, and 3rd interwave subperiods, respectively). Similar trends of higher risks of death during the wave subperiods were found in the matched-control individuals. In contrast, the 1^st^ and 2^nd^ interwave subperiods were associated with a lower risk of death among control group (HR at 0.87 [0.82–0.93], 0.91 [0.87–0.96], and 0.97 [0.93– 1.02] for 1st, 2nd, and 3rd interwave subperiods, respectively).

**Table. 3.** Multivariable analysis of mortality in the 71,583 CDR and 143,166 matched-control individuals of the study during the prepandemic and pandemic periods (cause-specific Cox models).

| Model | Variable | CDR |  |  | Matched controls individuals |  |  |
| --- | --- | --- | --- | --- | --- | --- | --- |
|  |  | Deaths observed (n) / person-years (n) | HR [95% CI] | p value | Deaths observed (n) / person-years (n) | HR [95% CI] | p value |
| Model 4 | Prepandemic period | 14,455 / 98,444 | 1 (reference) |  | 10,898 / 231,672 | 1 (reference) |  |
|  | Pandemic period | 13,932 / 86,465 | 1.08 [1.05–1.10] | <0.01 | 11,387 / 222,760 | 1.05 [1.02–1.08] | <0.01 |
| Model 5 | Prepandemic period | 14,021 / 98,444 | 1 (reference) |  | 10,898 / 231,672 | 1 (reference) |  |
|  | Pandemic period, no history of COVID-19-related hospitalization | 11,924 / 82,804 | 0.98 [0.95–1.00] | 0.07 | 10,110 / 220,149 | 0.96 [0.93–0.98] | <0.01 |
|  | Pandemic period, ≥1 COVID-19-related hospitalization | 2,008 / 3,661 | 3.48 [3.32–3.65] | <0.01 | 1,277 / 2,611 | 7.54 [7.11–8.00] | <0.01 |
| Model 6 | Prepandemic period | 14,455 / 98,444 | 1 (reference) |  | 10,898 / 231,672 | 1 (reference) |  |
|  | 1<sup>st</sup> wave | 1,235 / 6,837 | 1.19 [1.13–1.27] | <0.01 | 1,036 / 17,386 | 1.22 [1.14–1.30] | <0.01 |
|  | 1<sup>st</sup> interwave | 1,391 / 9,892 | 0.93 [0.88–0.97] | 0.01 | 1,075 / 25,117 | 0.87 [0.82–0.93] | <0.01 |
|  | 2<sup>nd</sup> wave | 3,860 / 21,604 | 1.19 [1.15–1.23] | <0.01 | 3,234 / 55,077 | 1.21 [1.15–1.25] | <0.01 |
|  | 2<sup>nd</sup> interwave | 2,552 / 17,503 | 0.98 [0.94–1.02] | 0.28 | 1,966 / 44,564 | 0.91 [0.87–0.96] | <0.01 |
|  | 3<sup>rd</sup> wave | 2,208 / 13,075 | 1.12 [1.07–1.17] | <0.01 | 1,843 / 33,764 | 1.13 [1.07–1.18] | <0.01 |
|  | 3<sup>rd</sup> interwave | 2,686 / 17,554 | 1.03 [0.98–1.07] | 0.24 | 2,233 / 46,852 | 0.97 [0.93–1.02] | 0.20 |

All HR shown correspond to cause-specific Cox regressions also adjusted for the following covariates: sex, age category, diabetes, and cardiovascular disease. Abbreviations used: CI, confidence interval; CDR, chronic dialysis recipients; HR, hazard ratio.

### 3.3 Vaccination

Figure 3 presents the dynamics of COVID-19 vaccination. By 31 December 2022, nearly 90% of CDR had received the two initial doses of vaccine, 82% had received a third dose, and 54% had received a fourth dose. This observed decrease in vaccination coverage over time was even greater in the matched-control individuals. In CDR, receiving two doses was associated with a 63% (HR = 0.37 [0.32– 0.42]) lower risk of COVID-19-related hospital admissions (Table 4), and a 9% (HR = 0.91 [0.86–0.97]) lower risk of all-cause mortality (Table 5). Receiving additional booster doses was associated with lower risks of COVID-19 related hospitalization (HR at 0.66 [0.56–0.77] and 0.83 [0.72–0.94] for additional third and fourth doses, respectively), and all-cause mortality (HR at 0.55 [0.51–0.59] and 0.88 [0.83– 0.95] for additional third and fourth doses, respectively). Almost similar results were found in matched-control individuals for COVID-19-related hospitalization (HR = 0.28 [0.24–0.32], 0.60 [0.51–0.72], 0.77 [0.62–0.98] for receiving additional second, third and fourth doses, respectively), and all-cause mortality (HR =0.70 [0.65–0.75], 0.61 [0.56–0.67], 0.81 [0.73–0.90], respectively).

**Figure 3.**
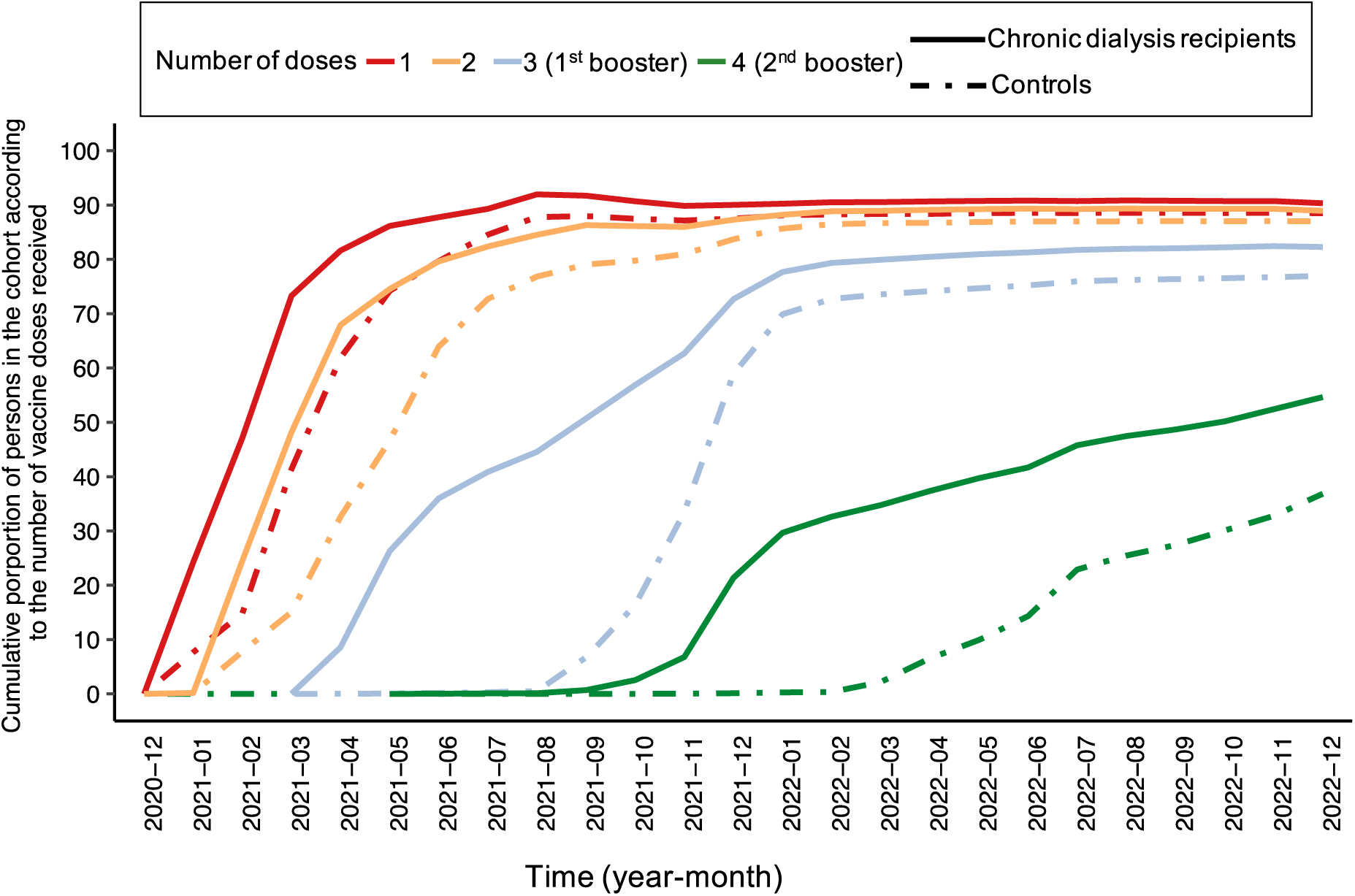
Dynamics of the COVID-19 vaccination in the chronic dialysis recipients included in the study and in the corresponding matched-control individuals.

**Table 4.** Multivariable analysis of COVID-19 hospitalization after a given number of vaccine doses received in the CDR and matched-control individuals (cause-specific Cox model).

| Number of vaccine doses | CDR<br>Individuals with a<br>COVID-19<br>hospitalization (n) /<br>person-years (n) | HR [95% CI] | p value | Matched controls<br>Individuals with a<br>COVID-19<br>hospitalization (n) /<br>person-years (n) | HR [95% CI] | p value |
| --- | --- | --- | --- | --- | --- | --- |
| 0 or 1 | 3,006 / 39,494 | 1 (reference) |  | 2,397 / 116,699 | 1 (reference) |  |
| 2 (vs. ≤ 1) | 351 / 12,149 | 0.37 [0.32–0.42] | <0.01 | 264 / 42,935 | 0.28 [0.24–0.32] | <0.01 |
| 3 (1<sup>st</sup> booster, vs. 2) | 618 / 19,430 | 0.66 [0.56–0.77] | <0.01 | 486 / 49,045 | 0.60 [0.51–0.72] | <0.01 |
| 4 (2<sup>nd</sup> booster, vs. 3) | 396 / 6,915 | 0.83 [0.72–0.94] | <0.01 | 113 / 8,554 | 0.77 [0.62–0.98] | 0.03 |

All HR shown correspond to cause-specific Cox regressions also adjusted for the following covariates: sex, age category, diabetes, cardiovascular disease, and wave and interwave subperiods; they are reported per additional vaccine dose (i.e. 2 vs. ≤ 1, 3 vs. 2, and 4 vs. 3, respectively). Abbreviations used: CI, confidence interval; CDR, chronic dialysis recipients; HR, hazard ratio.

**Table 5.** Multivariable analysis of survival after a given number of vaccine doses received in the CDR and matched-control individuals (cause-specific Cox model).

| Number of vaccine doses | CDR<br>Deaths observed (n) /<br>person-years (n) | HR [95% CI] | p value | Matched controls<br>Deaths observed (n) /<br>person-years (n) | HR [95% CI] | p value |
| --- | --- | --- | --- | --- | --- | --- |
| 0 or 1 | 7,137 / 41,084 | 1 (reference) |  | 6,651 / 118,095 | 1 (reference) |  |
| 2 (vs. ≤ 1) | 2,246 / 12,960 | 0.91 [0.86–0.97] | <0.01 | 1,949 / 43,567 | 0.70 [0.65–0.75] | <0.01 |
| 3 (1<sup>st</sup> booster, vs. 2) | 2,683 / 20,320 | 0.55 [0.51–0.59] | <0.01 | 2,174 / 49,501 | 0.61 [0.56–0.67] | <0.01 |
| 4 (2<sup>nd</sup> booster, vs. 3) | 1,638 / 10,548 | 0.88 [0.83–0.95] | <0.01 | 581 / 11,159 | 0.81 [0.73–0.90] | <0.01 |

All HR shown correspond to cause-specific Cox regressions also adjusted for the following covariates: sex, age category, diabetes, cardiovascular disease, and wave and interwave subperiods; they are reported per additional vaccine dose (i.e. 2 vs. ≤ 1, 3 vs. 2, and 4 vs. 3, respectively). Abbreviations used: CI, confidence interval; CDR, chronic dialysis recipients; HR, hazard ratio.

### 3.4 Sensitivity Analyses

Sensitivity analyses using models based on the Fine and Gray model to handle competing risks (Tables S2 to S5 in the Supplementary Appendix) globally yielded similar results to those relying on cause-specific Cox models (Tables 2 to 5).

## 4. Discussion

This French national study based on the SNDS indicates that, compared with the prepandemic period, the likelihood of receiving a kidney transplant in CDR was lower during the pandemic period, especially during the initial wave. In addition, this likelihood was dramatically decreased in CDR with a history of COVID-19-related hospitalization. The higher risk of death observed during wave subperiods mainly concerned individuals with a history of COVID-19-related hospitalization. Importantly, similar mortality rates were observed in CDR and matched-control individuals. Vaccine booster coverage declined with additional doses, but receiving a higher number of doses was associated with lower risks of death and COVID-19-related hospitalization in both CDR and controls. The significant findings of the study are discussed below and organized according to three main points: transplantation, mortality, and vaccination.

### 4.1 Transplantation

Numerous studies have reported a decline in transplantation activity during the pandemic period compared with the prepandemic period.^23^ However, the evolving implications of this decline on the likelihood of CDR to receive a kidney transplant during the pandemic subperiods remain largely unexplored. To our knowledge, the present national study, which is based on real-world data is the first to have taken into account time-dependent covariates and the competitng risks between transplantation and death events, for assessing the impact of the pandemic on the likelihood of CDR to receive a transplant. The study indicates that, considering individuals with equal times since ESKD incidence date, the pandemic period was associated with a lower HR for receiving a kidney transplant as compared to the prepandemic period. The lower likelihood to receive a transplant during the pandemic was observed in CDR without a history of COVID-19-related hospitalization (Table 2, model 2), highlights an indirect impact of the pandemic. However, CDR with a history of COVID-19-related hospitalization had an additional decrease in the likelihood to receive a kidney transplant, suggesting an additional direct impact of the pandemic relating to long-term consequences of COVID-19 in CDR. Our study further indicates that all wave and interwave subperiods were associated with a lower likelihood to receive a transplant (Table 2, model 3).

### 4.2 Mortality

Our study suggests that despite the challenges faced by the French healthcare system during the pandemic, the survival of CDR without a history of COVID-19-related hospitalization was not impacted (see Model 5 in Table 3, HR = 0.98 [0.95–1.00]). In contrast, a dramatic higher risk of mortality was observed in those CDR and matched-control individuals who experienced COVID-19-related hospitalizations (see the stratified Cox model 5 in Table 3, HR = 3.48 [3.32–3.65] in CDR, and HR = 7.54 [7.11–8.00] in matched controls). The higher additional risk in matched controls than that in CDR, likely reflect that the incremental risk relating to COVID-19-related hospitalization in CDR is somewhat limited because their baseline risk of death is already substantially higher than that of the matched-control individuals (HR = 3.15 [3.07–3.23], see Table S1 in the Supplementary Appendix).

Previous studies in the USA and the UK have reported that mortality in CDR observed during the first wave of the COVID-19 pandemic was nearly 30% higher than the corresponding mortality trends considering the previous 5 years.^24,25^ The present study also estimates a 19% higher risk of death for CDR in France during the first wave (Model 6 in Table 3), and additionally extended analyses until December 31, 2022, exploring the evolving pattern of CDR mortality over wave and interwave pandemic subperiods. Study results (Model 6 in Table 3) indicate in particular that the excess risk of death remained substantial during the 2^nd^ and 3^rd^ waves, with a corresponding HR at 1.19 [1.15–1.23], and 1.12 [1.07–1.17], respectively. It is important to note that similar higher risks of death were also observed in matched-control individuals during all wave subperiods. Therefore, such higher risks were not specific of the corresponding kidney disease in the CDR under study, and continued well after the first wave in both vulnerable populations (CDR and matched controls).

### 4.3 Vaccination

Prior to the roll-out of COVID-19 vaccinations in France, studies found that hesitancy towards the COVID-19 vaccine in the general population reached nearly 30%.^26,27^ However, our investigations on the dynamics of COVID-19 vaccination coverage indicate that almost 90% of CDR received two doses of the COVID-19 vaccine (Figure 3). Above all, this high percentage reflects the fact that vaccination policy in France for the whole population, considering the two first doses, was highly recommended. The study analyses on additional (i.e., booster) doses deserve particular attention. Several studies have reported an association between COVID-19 vaccination and lower risks of COVID-19-related hospitalization or death,^6,7^ but due to waning vaccine immunity over time, researchers have emphasized the need for additional booster campaigns, particularly for vulnerable populations such as CDR. To date, only Alobaidi et al.^9^ reported that 22% of hemodialysis patients had reluctance to take up the COVID-19 booster vaccine. Our study indicates that booster campaigns did not achieve the same level of coverage as that observed for the two initial doses, with even worse coverage in the matched-control group. Despite the hesitancy or reluctance of some CDR to receive booster doses, the present study indicates a positive association between receiving increased doses of COVID-19 vaccine and lower risks of COVID-19-related hospitalization and death. To date, only one study conducted by Cohen-Hagai et al,^28^ based on the data from 1030 patients compared the clinical efficacy of receiving a fourth dose of vaccine versus three doses in dialysis patients, with a corresponding 44% lower risk of COVID-19-related mortality. In our study, receiving a fourth dose versus three doses was associated with a 12% lower risk of all-cause mortality, and a 24% lower risk of COVID-19-related hospitalization. Lastly, the present study evaluated the effectiveness of the COVID-19 vaccine in CDR from January 1, 2021, to December 31, 2022. Therefore, the study results also address the concerns raised by Rouphael and Bausch-Jurken regarding the lack of reports on vaccine effectiveness after January 2022, when the Omicron variant became predominant among dialysis recipients.^6^

### 4.4 Limitations and strengths of the study

Our study shares many limitations with those from a recent study that investigated the impact of the COVID-19 pandemic on persons living with a kidney transplant.^19^ The first limitation relies on SNDS data which do not include detailed medical/clinical and individual socioeconomic data. The first incidence date considered was January 1, 2015, and in consequence, the CDR under study were relatively recent ones. Therefore, the study results cannot be extended to CDR having spent more than 5 years under dialysis. However, the limitation on relatively recent incidence dates was a requirement, and allowed us to include persons for which the beginning of the disease and potential comorbidities were documented in the SNDS. Another limitation of the study arises from its observational nature. We attempted to address this limitation by adjusting estimates on the basis of comorbidities identified by the G10 mapping in SNDS data. The use of COVID-19-related hospitalizations as a proxy for severe COVID-19 cases could also lead to an underestimation of the direct impact of COVID-19. Factors such as variants, virus exposure, mitigation measures, changes in the health care system, vaccination schedules and unidentified confounding factors prevent from a straight-forward generalizability of our findings to other countries. However, similar drawbacks usually stand for any observational study. Nevertheless, our methodological framework could be used to plan comparable investigations in other countries, and even consider other diseases. Importantly, though this study is among the first ones to follow CDR almost two years after the beginning on the pandemic, this hindsight still remains too limited to identify long-term consequences of the pandemic in this population, which should be documented by further studies. The study has many strengths related to the methodological features of its experimental analysis plan, with four particular elements that are worth recalling. First, the results presented were obtained with a real-world evidence approach based on exhaustive nationwide data. Second, outcomes during the prepandemic and pandemic periods were compared considering individuals with similar times elapsed since ESKD incidence. Third, outcomes estimated in CDR were also estimated in matched-control individuals, enabling a comprehensive interpretation of the kidney disease *per se* contribution in the CDR estimates. Fourth, the results from analyses based on the Fine and Gray model to handle competing risks (see sensitivity analyses in the Supplementary Appendix) were roughly similar to those based on cause-specific Cox regressions presented as the main analyses, and therefore, the evolving patterns reported in the present study appear particularly robust.

In conclusion, this national study details the dynamics of kidney transplantation access, survival, and vaccination over the COVID-19 pandemic subperiods in CDR having an incident date within years 2015 to 2021. Considering the pandemic period as a whole, we did not observe a higher mortality among CDR without a history of COVID-19-related hospitalization, suggesting that efforts to limit the indirect impact of the pandemic on survival in this vulnerable population have been successful. During the three wave subperiods, similar excess mortality rates were observed both in CDR and matched-control individuals, indicating that this excess mortality was more related to contextual covariates (e.g., comorbidities, age, …) than to the kidney disease, strictly speaking. Vaccine coverage decreased with each additional booster dose, although receiving additional booster doses was associated with significant benefits. Therefore, study results argue in favor of future research into candidate policies aimed at increasing adherence to vaccine booster doses.

## Acknowledgements

The COVID-HOSP working group: Tristan Delory, Centre Hospitalier Annecy Genevois, Annecy, France; Fanny Duchaine, IRDES, Paris, France; Maude Espagnacq, IRDES, Paris, France; Gilles Hejblum, INSERM, Paris, France; Myriam Khlat, INED, Aubervilliers, France; Nathanaël Lapidus, INSERM, Paris, France; Sophie Le Cœur, INED, Aubervilliers, France; Elhadji Leye, INSERM, Paris, France; Paul Moulaire, INSERM, Paris FRANCE; Jonas Poucineau, INED, Aubervilliers, France.

## Data availability statement

According to the principles of data protection and French regulations, the authors cannot publicly release the data from the French National Health Data System (SNDS). However, any person or structure, public or private, for-profit or non-profit, can access SNDS data upon authorization from the French Data Protection Office (CNIL, Commission Nationale de l’Informatique et des Libertés) to carry out a study, research, or an evaluation of public interest (https://www.snds.gouv.fr/SNDS/Contexte-et-perspectives-reglementaires#).

## Author Contributions

TD, GH, and NL initiated the study. GH and NL supervised the study; KEK, GH, NL, and EL designed the experimental plan; EL managed data and performed the analyses; GH, NL, and EL can take responsibility for the integrity of the data and the accuracy of the data analysis, EL is the guarantor; GH, NL, and EL prepared the first draft of the manuscript; All authors (TD, ME, KEK, GH, MK, NL, SLC, and EL) contributed to interpretation of the data, critically revised the manuscript, and approved the final version.

