## supplementary material for "Evolving impact of the COVID-19 pandemic in chronic dialysis recipients over the course of pandemic waves and COVID-19 vaccination rollout: a French national study"

### Supplementary material to "Assessing the evolving impact of the COVID-19 pandemic in chronic dialysis recipients along the wave subperiods and vaccination rollout: a French national study" by Leye E et al.

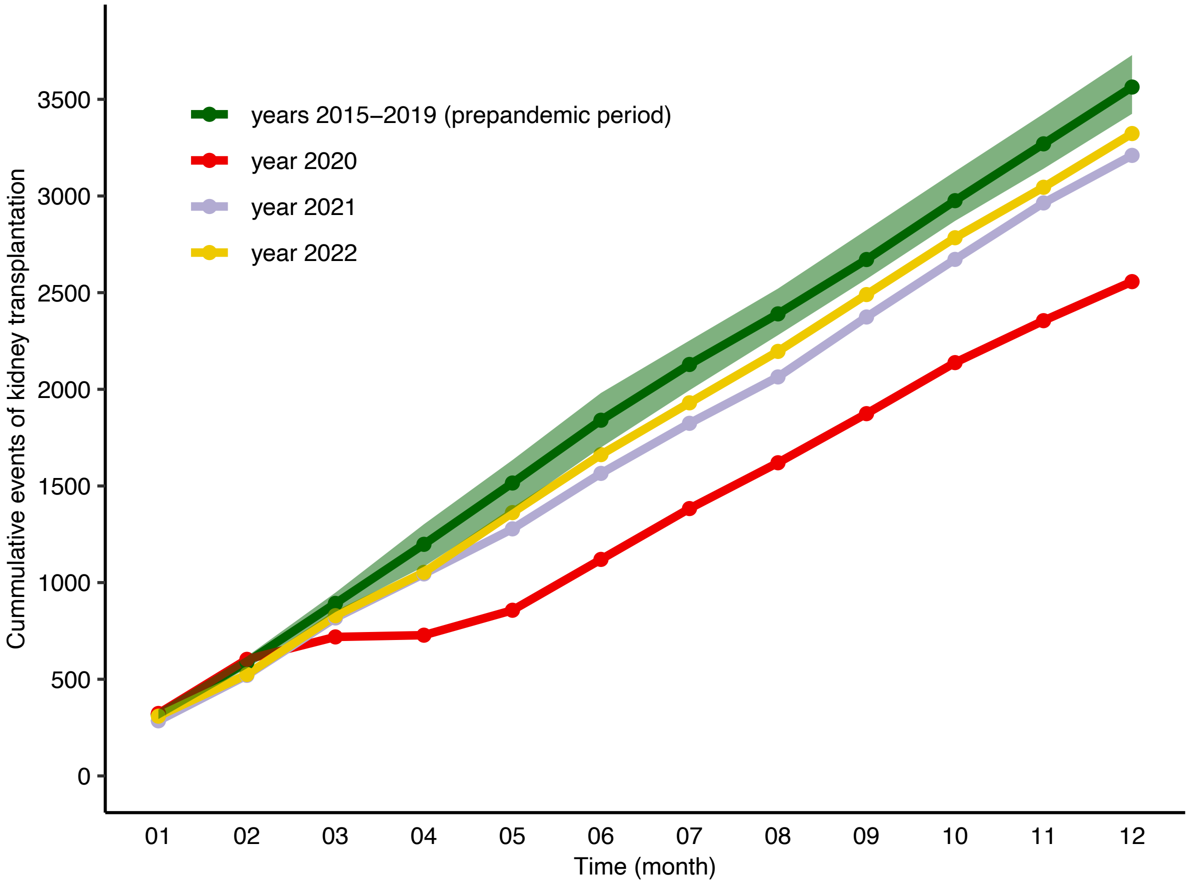


**Supplementary Figure S1: Kidney transplantation events among CDR in France in years 2015 to 2022.**
Prepandemic years (years 2015 to 2019) were pooled, and dark green solid line indicates the cumulative mean value observed each month considering the 5 corresponding years, while the light green area delineates the minimum and maximum observed range.

**Supplementary Table S1. Multivariable analysis of mortality in the 71,583 chronic dialysis patients and 143,166 matched controls during the prepandemic and pandemic periods.**

| Variable | Variable value | Deaths observed (n) / person-years (n) | HR [95% CI] | p value |
|---|---|---|---|---|
| Period | Prepandemic | 25,353 / 330,116 | 1 (reference) | |
| | Pandemic | 25,319 / 309225 | 1.03 [1.00–1.06] | 0.02 |
| Population | Matched-control individuals | 22,285 / 454,432 | 1 (reference) | |
| | CDR | 28,387 / 184,909 | 3.15 [3.07–3.23] | <0.01 |
| interaction | Period × population | | 1.02 [0.98–1.05] | 0.34 |

Abbreviations used: CI, confidence interval; HR, hazard ratio.

**Supplementary Table S2. Multivariable analysis of kidney transplantation events in the 71,583 chronic dialysis recipients of the study during the prepandemic and pandemic periods (Fine and Gray models).**

| Model | Variable | Kidney transplantations (n) / person-years (n) | SHR [95% CI] | p value |
|---|---|---|---|---|
| Model S1 | Prepandemic period | 5,853 / 98,444 | 1 (reference) | |
| | Pandemic period | 4,172 / 86,465 | 0.80 [0.77–0.83] | <0.01 |
| Model S2 | Prepandemic period | 5,853 / 98,444 | 1 (reference) | |
| | Pandemic period, no history of COVID-19-related hospitalization | 4,060 / 82,804 | 0.81 [0.78–0.85] | <0.01 |
| | Pandemic period, ≥1 COVID-19-related hospitalization | 112 / 3,661 | 0.56 [0.47–0.68] | <0.01 |
| Model S3 | Prepandemic period | 5,853 / 98,444 | 1 (reference) | |
| | 1st wave | 67 / 6,837 | 0.17 [0.13–0.21] | <0.01 |
| | 1st interwave | 505 / 9,892 | 0.88 [0.81–0.97] | <0.01 |
| | 2nd wave | 1,024 / 21,604 | 0.79 [0.74–0.85] | <0.01 |
| | 2nd interwave | 874 / 17,503 | 0.84 [0.78–0.90] | <0.01 |
| | 3rd wave | 673 / 13,075 | 0.83 [0.77–0.90] | <0.01 |
| | 3rd interwave | 1,029 / 17,553 | 0.94 [0.88–1.00] | >0.01 |

All SHR shown correspond to Fine and Gray regression models also adjusted for the following covariates: sex, age category, diabetes, and cardiovascular disease.
Abbreviations used: CI, confidence interval; SHR, subdistribution hazard ratio.

**Supplementary Table S3. Multivariable analysis of mortality in the 71,583 chronic dialysis recipients and 143,166 matched-control individuals of the study during the prepandemic and pandemic periods (Fine and Gray models).**

| Model | Variable | Chronic dialysis recipients Deaths observed (n) / person-years (n) | SHR [95% CI] | p value | Matched-controls individuals Deaths observed (n) / person-years (n) | SHR [95% CI] | p value |
|---|---|---|---|---|---|---|---|
| Model S4 | Prepandemic period | 14,455 / 98,444 | 1 (reference) | | 10,898 / 231,672 | 1 (reference) | |
| | Pandemic period | 13,932 / 86,465 | 1.14 [1.11–1.17] | <0.01 | 11,387 / 222,760 | 1.08 [1.05–1.11] | <0.01 |
| Model S5 | Prepandemic period | 14,021 / 98,444 | 1 (reference) | | 10,898 / 231,672 | 1 (reference) | |
| | Pandemic period, no history of COVID-19-related hospitalization | 11,924 / 82,804 | 1.03 [1.01–1.06] | 0.01 | 10,110 / 220,149 | 0.99 [0.95–1.01] | 0.31 |
| | Pandemic period, ≥1 COVID-19-related hospitalization | 2,008 / 3,661 | 3.56 [3.38–3.74] | <0.01 | 1,277 / 2,611 | 7.58 [7.11–8.08] | <0.01 |
| Model S6 | Prepandemic period | 14,455 / 98,444 | 1 (reference) | | 10,898 / 231,672 | 1 (reference) | |
| | 1st wave | 1,235 / 6,837 | 1.30 [1.22–1.37] | <0.01 | 1,036 / 17,386 | 1.27 [1.19–1.35] | <0.01 |
| | 1st interwave | 1,391 / 9,892 | 0.98 [0.92–1.03] | 0.41 | 1,075 / 25,117 | 0.90 [0.84–0.95] | <0.01 |
| | 2nd wave | 3,860 / 21,604 | 1.26 [1.22–1.31] | <0.01 | 3,234 / 55,077 | 1.24 [1.19–1.29] | <0.01 |
| | 2nd interwave | 2,552 / 17,503 | 1.03 [0.98–1.07] | 0.22 | 1,966 / 44,564 | 0.93 [0.89–0.98] | <0.01 |
| | 3rd wave | 2,208 / 13,075 | 1.18 [1.13–1.23] | <0.01 | 1,843 / 33,764 | 1.16 [1.10–1.22] | <0.01 |
| | 3rd interwave | 2,686 / 17,554 | 1.10 [1.05–1.14] | <0.01 | 2,233 / 46,852 | 1.00 [0.96–1.05] | 0.93 |

All SHR shown correspond to Fine and Gray regression models also adjusted for the following covariates: sex, age category, diabetes, and cardiovascular disease. Abbreviations used: CI, confidence interval; SHR, subdistribution hazard ratio.

**Supplementary Table S4. Multivariable analysis of COVID-19 hospitalization after a given number of vaccine doses received in the chronic dialysis recipients and matched-control individuals (Fine and Gray model).**

| Number of vaccine doses | Chronic dialysis recipients Persons with a COVID-19 hospitalization (n) / person-years (n) | SHR [95% CI] | p value | Matched-controls individuals Persons with a COVID-19 hospitalization (n) / person-years (n) | SHR [95% CI] | p value |
|---|---|---|---|---|---|---|
| 0 or 1 | 3,006 / 39,494 | 1 (reference) | | 2,397 / 116,699 | 1 (reference) | |
| 2 (vs. ≤ 1) | 351 / 12,149 | 0.36 [0.32–0.41] | <0.01 | 264 / 42,935 | 0.29 [0.25–0.33] | <0.01 |
| 3 (1st booster, vs. 2) | 618 / 19,430 | 0.66 [0.58–0.76] | <0.01 | 486 / 49,045 | 0.68 [0.57–0.80] | <0.01 |
| 4 (2nd booster, vs. 3) | 396 / 6,915 | 0.76 [0.68–0.87] | <0.01 | 113 / 8,554 | 0.82 [0.65–1.03] | 0.08 |

All SHR shown correspond to Fine and Gray regression models also adjusted for the following covariates: sex, age category, diabetes, cardiovascular disease, and waves and interwaves subperiods; they are reported per additional vaccine dose (i.e. 2 vs. ≤ 1, 3 vs. 2, and 4 vs. 3, respectively).
Abbreviations used: CI, confidence interval; SHR, subdistribution hazard ratio.

**Supplementary Table S5. Multivariable analysis of survival after a given number of vaccine doses received in the chronic dialysis recipients and matched-control individuals (Fine and Gray model).**

| Number of vaccine doses | Chronic dialysis recipients<br>Deaths observed (n) / person-years (n) | SHR [95% CI] | p value | Matched-controls individuals<br>Deaths observed (n) / person-years (n) | SHR [95% CI] | p value |
|---|---|---|---|---|---|---|
| 0 or 1 | 7,137 / 41,084 | 1 (reference) | | 6,651 / 118,095 | 1 (reference) | |
| 2 (vs. ≤ 1) | 2,246 / 12,960 | 0.81 [0.77–0.86] | <0.01 | 1,949 / 43,567 | 0.65 [0.61–0.69] | <0.01 |
| 3 (1st booster, vs. 2) | 2,683 / 20,320 | 0.56 [0.53–0.60] | <0.01 | 2,174 / 49,501 | 0.61 [0.53–0.66] | <0.01 |
| 4 (2nd booster, vs. 3) | 1,638 / 10,548 | 0.88 [0.82–0.94] | <0.01 | 581 / 11,159 | 0.80 [0.73–0.89] | <0.01 |

All SHR shown correspond to Fine and Gray regression models also adjusted for the following covariates: sex, age category, diabetes, cardiovascular disease, and waves and interwaves subperiods; they are reported per additional vaccine dose (i.e. 2 vs. ≤ 1, 3 vs. 2, and 4 vs. 3, respectively).
Abbreviations used: CI, confidence interval; SHR, subdistribution hazard ratio.
